# Associations of Epstein-Barr Virus Infection with Attention Deficit Hyperactivity Disorder, Learning Disability, and Special Education in U.S. Children

**DOI:** 10.1101/2021.12.09.21267546

**Authors:** Jingjing Wang, Yaping Li, Xiaozhen Geng, Xin Zhang, Yanfeng Xiao, Wenjun Wang

## Abstract

**Background:** Most infections of Epstein-Barr virus (EBV), which is potentially neurotropic, occurred in childhood, but little is known about its association with child neurodevelopmental outcomes.

**Methods:** We determined whether EBV seropositivity was associated with parent-reported attention deficit hyperactivity disorder (ADHD), learning disability, or special education utilization among children, using data from the National Health and Nutrition Examination Survey (NHANES) 2003-2004. Potential confounding factors were adjusted using survey logistic regression models.

**Results:** EBV seroprevalence was 69.6% (95% CI, 67.1%-72.1%) for U.S. children aged 6-19. The prevalence was 8.86% (95% CI, 7.47%-10.47%) for ADHD among 6-19 year olds, 11.70% (95% CI, 9.84%-13.87%) for learning disability among 6-15 year olds, and 10.18% (95% CI, 8.58%-12.05%) for special education among 6-17 year olds. Children with positive anti-EBV had higher crude prevalence rates of learning disability and special education but not ADHD compared with those with negative anti-EBV. The adjusted odds ratios were 2.76 (95% CI, 1.53-4.96) for learning disability, 3.58 (95% CI, 1.92-6.55) for special education, and 0.77 (95% CI, 0.42-1.38) for ADHD, when comparing children with positive and negative anti-EBV.

**Conclusions:** EBV seropositivity was associated with learning disability and special education among U.S. children. Future studies that longitudinally examine the associations are warranted.

## 1. Introduction

Infection with Epstein-Barr virus (EBV), a member of the herpesvirus family, is very common worldwide. The infection usually occurs during childhood and adolescence. By adulthood, up to 90% individuals have been infected and have antibodies against the virus (1). EBV infection has a wide range of disease spectrum (2). Most infections go unnoticed or produce very mild symptoms. In some cases, EBV causes infectious mononucleosis, which is usually self-limited but occasionally be life-threatening due to serious complications, especially complications of central nervous system (CNS) (3-5). What’s more, EBV was identified as the first oncogenic virus to human cancer, and has been linked to Burkitt’ lymphoma, B cell lymphoma, Hodgkin’s diseases, nasopharyngeal carcinoma, and gastric carcinoma (6). Its associations with autoimmune diseases have also been reported (7).

Neurodevelopmental and learning problems in children are of great concern and infection is considered as one of the major contributors (8). Although the effects of maternal or congenital infections are more often described, studies showed that infections in early to middle childhood can also lead to neurodevelopmental impairment.(8) EBV is potentially neurotropic and most infections of EBV occur in childhood (5, 9). Thus, it would be interesting to investigate whether EBV infection affects child neurodevelopmental outcomes. Involved studies usually focused on EBV infection and cognitive functioning, but their results are controversial (10-12). Studies of EBV and learning problems are lacking.

In this study, we analyzed a large representative sample of U.S. 6-19 years of age from the National Health and Nutrition Examination Survey (NHANES), to determine whether EBV seropositivity was associated with reported attention deficit hyperactivity disorder (ADHD), learning disability, or special education utilization.

## 2 Materials and Methods

### 2.1 Study population

Data were from NHANES 2003-2004. The NHANES is a population-based survey, conducted by the National Center for Health Statistics (NCHS) to obtain a representative sample of the noninstitutionalized civilian population of the United States. The survey was based on a complex, multi-stage, stratified probability cluster design. It collects information every two years on health and nutrition of the U.S. household population. Detailed survey procedures and consent documents of the NHANES 2003-2004 are available on the NCHS website (https://wwwn.cdc.gov/nchs/nhanes/Default.aspx).

In NHANES 2003-2004, EBV antibody testing was conducted among participants aged 6-19 years at the time of survey interview. Of the 3,337 participants aged 6-19 years in the survey, 2,870 had enough stored serum samples to test EBV antibody. Among these 2,870 participants, 2,849 had interpretable data on EBV infection. After excluding 2 individuals without any data on neurodevelopmental outcomes, the remaining 2,847 participants constitute the study sample. In NHANES 2003-2004, ADHD was evaluated in participants aged 4-19 years, learning disability in participants aged 4-15 years, and special education utilization in participants aged 1-17 years. Thus, among the 2,847 included participants, 2,844 had available data for ADHD (aged 6-19 years), 1,818 for learning disability (aged 6-15 years), and 2,339 for special education utilization (aged 6-17 years) (Figure 1). The study was exempted from institutional review board approval through Second Affiliated Hospital of Xi’an Jiaotong University. The report followed the Strengthening the Reporting of Observational Studies in Epidemiology (STROBE) guideline.

**Figure 1.**
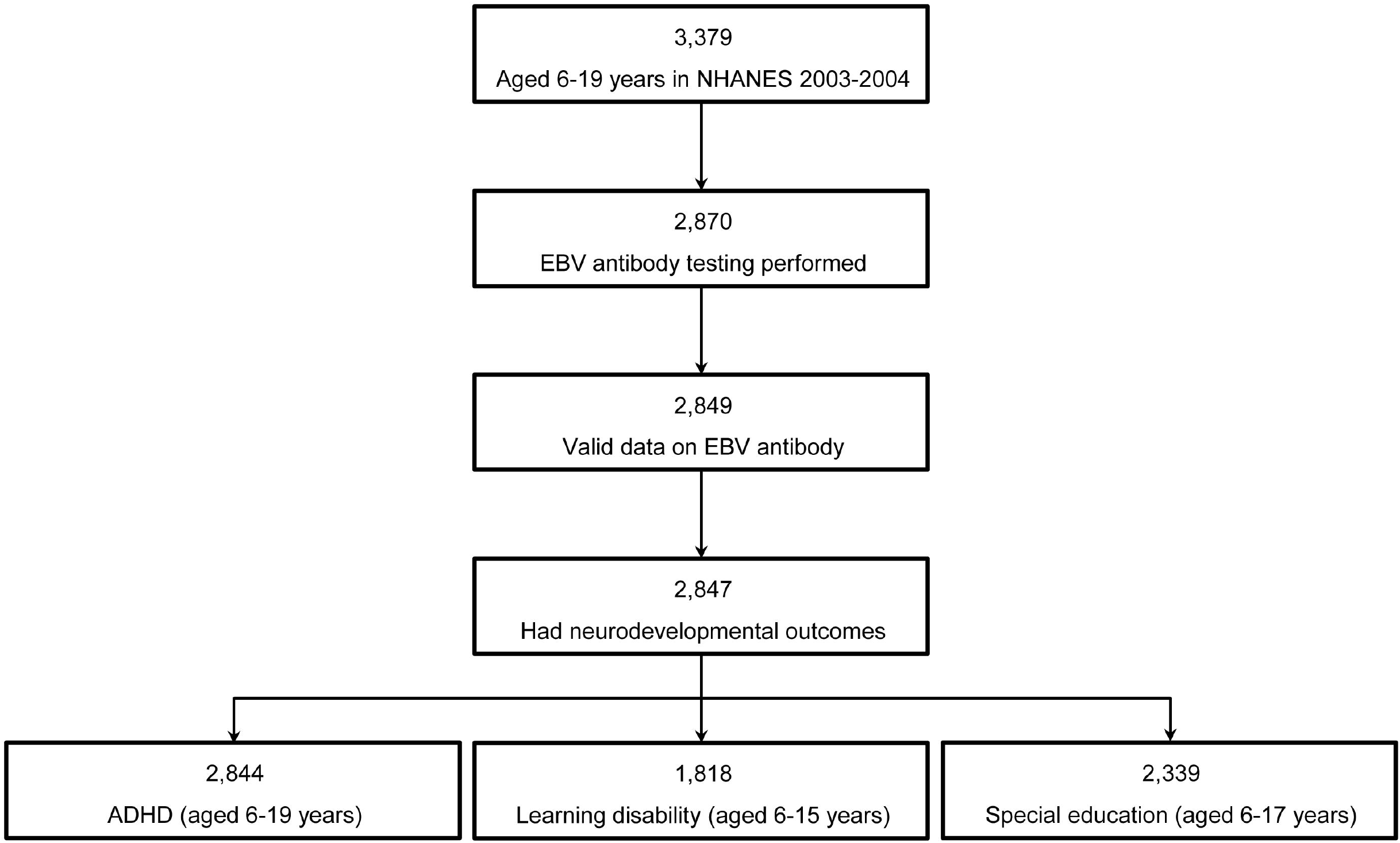
Determination of the Study Sample. ADHD, attention deficit hyperactivity disorder.

**Figure 2.**
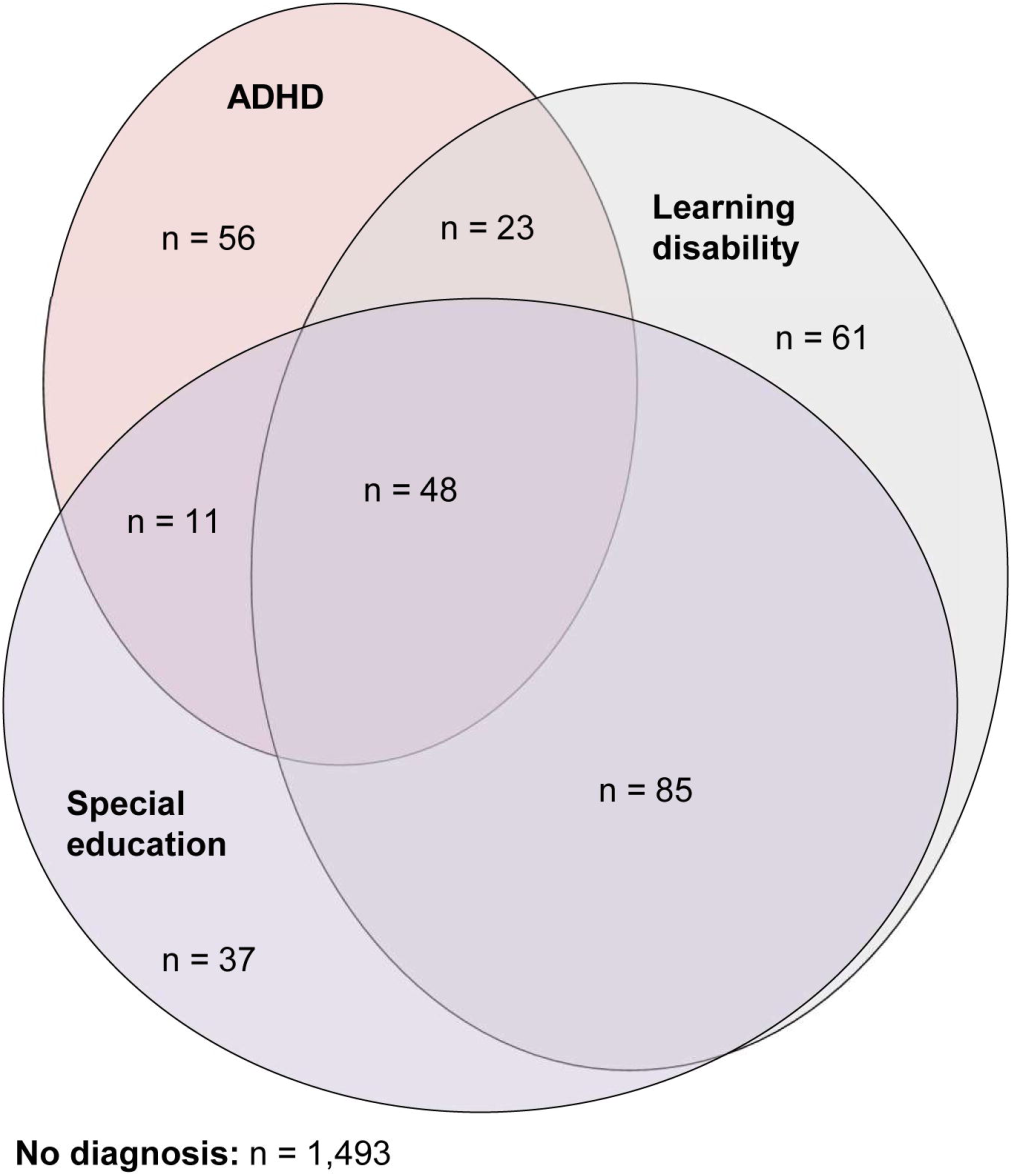
The Co-occurrence of the Neurodevelopmental Outcomes among the 1,814 Participants with Information on All Three Outcomes. ADHD, attention deficit hyperactivity disorder.

### 2.2 EBV antibody testing

Immunoglobulin (IgG) antibody against EBV viral capsid antigen was measured using a commercial enzyme immunoassay kit (Diamedix, Miami, FL). The sensitivity of the assay is 96.6% and the specificity is 97.7%. All QA/QC procedures recommended by the manufacturer were followed. Detailed description of laboratory methodology can be accessed on the NCHS website (https://wwwn.cdc.gov/Nchs/Nhanes/2003-2004/SSEBV_C.htm). Data were recorded as positive (EIA index ≥ 1.10), negative (EIA index < 0.90), or equivocal (EIA index ≥ 0.90 to 1.09). Since only a small fraction of the subjects (∼0.05%) were recorded as equivocal, they were excluded from our analysis.

### 2.3 Neurodevelopmental outcomes

Neurodevelopmental outcomes included reported ADHD, learning disability, and special education utilization. The assessment was based on parental/guardian response. For ADHD assessment, the participants were asked “has a doctor or health professional ever told you/survey participant (SP) that you/s/he/SP had attention deficit disorder?” For learning disability assessment, the participants were asked “has a representative from a school or a health professional ever told you/SP that s/he/SP had a learning disability?” For the assessment of special education utilization, the question was as follows: “does SP receive special education or early intervention services?”

### 2.4 Confounding variables

In addition to EBV infection and reported neurodevelopmental outcomes, NHANES 2003-2004 collected information on other covariates (13, 14). Race/ethnicity was divided into four categories: non-Hispanic white, non-Hispanic black, Mexican American, and other. Educational level of the household reference person was divided into three categories: less than high school, high school completion or equivalent, and more than high school. Poverty level was evaluated using a poverty income ratio, which was based on self-report of family income, family size, and tables published annually by the U.S. Census Bureau. Health insurance status of the participants was divided as insurance covered or not. Household size was coded as ≤ 4 or > 4. Whether there was a smoker in the home (yes/no) was used to evaluate smoke exposure of the included participants. Information about the participants’ early life included whether the child’s mother smoked during pregnancy (yes/no), mother’s age at child’s birth (< 25, 25-35, and ≥ 35), low birth weight (< 2,500 g), receipt of any newborn care (e.g., in an intensive care unit, premature nursery, or any other type of special care facility), and attendance of day care or preschool.

### 2.5 Statistical analysis

The overall and characteristic-specific EBV seroprevalences were presented as percentages and 95% confidence intervals (CIs). EBV seroprevalences by each participant characteristic were compared using the Rao-Scott chi-square tests, corrected for survey design. To assess associations between EBV infection and developmental outcomes (ADHD, learning disability, and special education utility), we used univariate and multivariate survey logistic regression models to estimate crude and adjusted odds ratios (ORs) and corresponding 95% CIs. For each developmental outcome, we constructed three models to assess its association with EBV infection: a) logistic regression models adjusted for age, sex, and race/ethnicity; b) logistic regression models further adjusted for socioeconomic factors, including poverty level, health insurance coverage, household size, smoker in the home, educational level of the household reference person; c) full models also adjusted for early life characteristics, including whether the child’s mother smoked during pregnancy, mother’s age at child’s birth, low birth weight, receipt of any newborn care, and attendance of day care or preschool.

Sampling weights were used in all analyses to provide accurate estimates that were representative of the U.S. population during the survey time. All analyses were conducted with Stata/SE 12.0 (StataCorp LP, College Station, TX). All tests were 2-sided, and *P* < 0.05 was considered statistically significant. Data analysis was conducted from October to December 2020.

## 3. Results

In overall, antibodies to EBV were present in 80.54% (2,293/2,847) of the study population. Accounting for the survey design, the estimated EBV seroprevalence was 69.6% (95% CI, 67.1%-72.1%) for U.S. children aged 6-19. Characteristics of the 2,847 participants and EBV seropositivity by participant characteristics are presented in Table 1. Characteristics associated with higher EBV seroprevalence were female gender, older age, Mexican-Americans and Non-Hispanic Blacks, lower education level of household and family income, not covered by health insurance, larger household size, household smoke exposure, born to younger mothers, and not attending preschool (Table 1).

**Table 1.**
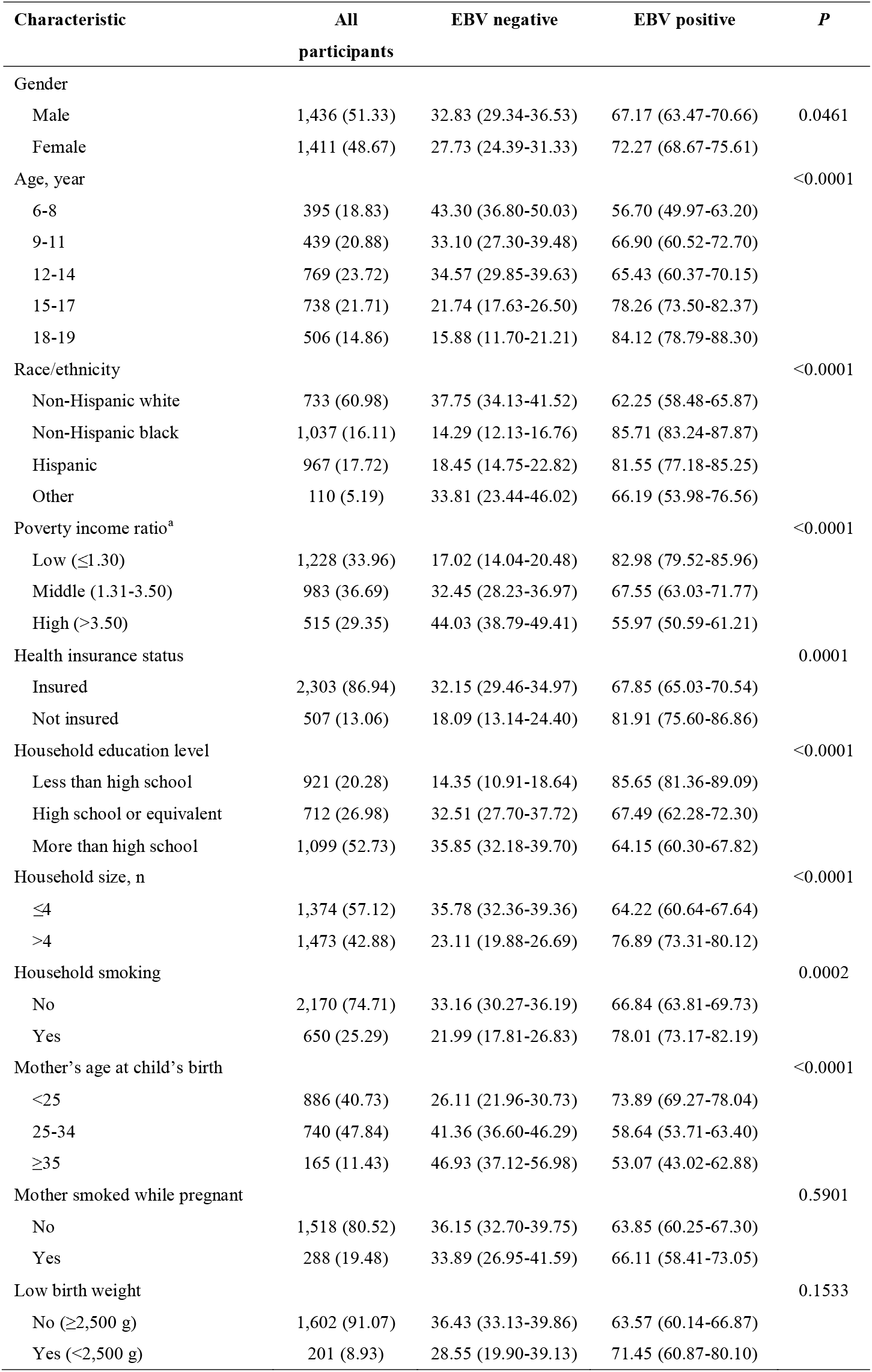
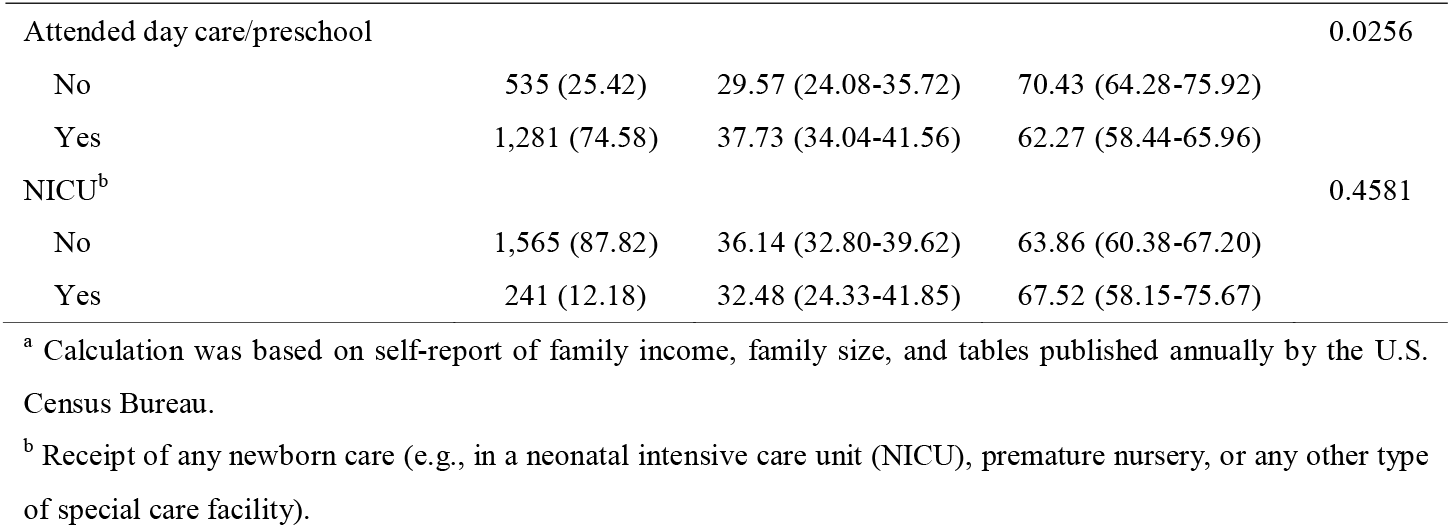
Characteristics of the 2,847 Participants Aged 6-19 and EBV Seroprevalence by Demographic Category

| Characteristic | All participants | EBV negative | EBV positive | P |
| --- | --- | --- | --- | --- |
| Gender |  |  |  |  |
| Male | 1,436 (51.33) | 32.83 (29.34-36.53) | 67.17 (63.47-70.66) | 0.0461 |
| Female | 1,411 (48.67) | 27.73 (24.39-31.33) | 72.27 (68.67-75.61) |  |
| Age, year |  |  |  | <0.0001 |
| 6-8 | 395 (18.83) | 43.30 (36.80-50.03) | 56.70 (49.97-63.20) |  |
| 9-11 | 439 (20.88) | 33.10 (27.30-39.48) | 66.90 (60.52-72.70) |  |
| 12-14 | 769 (23.72) | 34.57 (29.85-39.63) | 65.43 (60.37-70.15) |  |
| 15-17 | 738 (21.71) | 21.74 (17.63-26.50) | 78.26 (73.50-82.37) |  |
| 18-19 | 506 (14.86) | 15.88 (11.70-21.21) | 84.12 (78.79-88.30) |  |
| Race/ethnicity |  |  |  | <0.0001 |
| Non-Hispanic white | 733 (60.98) | 37.75 (34.13-41.52) | 62.25 (58.48-65.87) |  |
| Non-Hispanic black | 1,037 (16.11) | 14.29 (12.13-16.76) | 85.71 (83.24-87.87) |  |
| Hispanic | 967 (17.72) | 18.45 (14.75-22.82) | 81.55 (77.18-85.25) |  |
| Other | 110 (5.19) | 33.81 (23.44-46.02) | 66.19 (53.98-76.56) |  |
| Poverty income ratio<sup>a</sup> |  |  |  | <0.0001 |
| Low ($\leq 1.30$) | 1,228 (33.96) | 17.02 (14.04-20.48) | 82.98 (79.52-85.96) | |
| Middle (1.31-3.50) | 983 (36.69) | 32.45 (28.23-36.97) | 67.55 (63.03-71.77) |  |
| High ($> 3.50$) | 515 (29.35) | 44.03 (38.79-49.41) | 55.97 (50.59-61.21) | |
| Health insurance status |  |  |  | 0.0001 |
| Insured | 2,303 (86.94) | 32.15 (29.46-34.97) | 67.85 (65.03-70.54) |  |
| Not insured | 507 (13.06) | 18.09 (13.14-24.40) | 81.91 (75.60-86.86) |  |
| Household education level |  |  |  | <0.0001 |
| Less than high school | 921 (20.28) | 14.35 (10.91-18.64) | 85.65 (81.36-89.09) |  |
| High school or equivalent | 712 (26.98) | 32.51 (27.70-37.72) | 67.49 (62.28-72.30) |  |
| More than high school | 1,099 (52.73) | 35.85 (32.18-39.70) | 64.15 (60.30-67.82) |  |
| Household size, n |  |  |  | <0.0001 |
| $\leq 4$ | 1,374 (57.12) | 35.78 (32.36-39.36) | 64.22 (60.64-67.64) | |
| $> 4$ | 1,473 (42.88) | 23.11 (19.88-26.69) | 76.89 (73.31-80.12) | |
| Household smoking |  |  |  | 0.0002 |
| No | 2,170 (74.71) | 33.16 (30.27-36.19) | 66.84 (63.81-69.73) |  |
| Yes | 650 (25.29) | 21.99 (17.81-26.83) | 78.01 (73.17-82.19) |  |
| Mother's age at child's birth |  |  |  | <0.0001 |
| $< 25$ | 886 (40.73) | 26.11 (21.96-30.73) | 73.89 (69.27-78.04) | |
| 25-34 | 740 (47.84) | 41.36 (36.60-46.29) | 58.64 (53.71-63.40) |  |
| $\geq 35$ | 165 (11.43) | 46.93 (37.12-56.98) | 53.07 (43.02-62.88) | |
| Mother smoked while pregnant |  |  |  | 0.5901 |
| No | 1,518 (80.52) | 36.15 (32.70-39.75) | 63.85 (60.25-67.30) |  |
| Yes | 288 (19.48) | 33.89 (26.95-41.59) | 66.11 (58.41-73.05) |  |
| Low birth weight |  |  |  | 0.1533 |
| No ($\geq 2,500$ g) | 1,602 (91.07) | 36.43 (33.13-39.86) | 63.57 (60.14-66.87) | |
| Yes ($< 2,500$ g) | 201 (8.93) | 28.55 (19.90-39.13) | 71.45 (60.87-80.10) | |
| EBV and Child Neurodevelopmental Outcomes |  |  |  |  |
| Attended day care/preschool |  |  |  | 0.0256 |
| No | 535 (25.42) | 29.57 (24.08-35.72) | 70.43 (64.28-75.92) |  |
| Yes | 1,281 (74.58) | 37.73 (34.04-41.56) | 62.27 (58.44-65.96) |  |
| NICU<sup>b</sup> |  |  |  | 0.4581 |
| No | 1,565 (87.82) | 36.14 (32.80-39.62) | 63.86 (60.38-67.20) |  |
| Yes | 241 (12.18) | 32.48 (24.33-41.85) | 67.52 (58.15-75.67) |  |

<sup>a</sup> Calculation was based on self-report of family income, family size, and tables published annually by the U.S. Census Bureau.
<sup>b</sup> Receipt of any newborn care (e.g., in a neonatal intensive care unit (NICU), premature nursery, or any other type of special care facility).

Parent-reported ADHD, learning disability, and special education were reported for 7.4% (209/2,844), 12.1% (219/1,818), and 9.8% (229/2,339) of the study population, respectively. The co-occurrence of the neurodevelopmental outcomes is presented in Figure 1. Accounting for the survey design, the estimated prevalence among U.S. children was 8.9% (95% CI, 7.5%-10.5%) for ADHD among 6-19 year olds, 11.7% (95% CI, 9.8%-13.9%) for learning disability among 6-15 year olds, and 10.2% (95% CI, 8.6%-12.1%) for special education among 6-17 year olds.

Children with positive anti-EBV had higher crude prevalence rates of parent-reported learning disability [14.9% (95% CI: 12.4%-17.9%) vs. 5.9% (95% CI: 3.9%-8.7%)] and special education [12.8% (95% CI: 10.7%-15.3%) vs. 4.8% (95% CI: 3.1%-7.4%)] but not ADHD [8.5% (95% CI: 7.0%-10.4%) vs. 9.7% (95% CI: 7.1%-13.1%)] compared with those with negative anti-EBV (Table 2). The unadjusted ORs were 2.82 (95% CI, 1.76-4.53) for learning disability, 2.90 (95% CI, 1.76-4.80) for special education, and 0.87 (95% CI, 0.58-1.31) for ADHD. After adjusting for potential confounding variables using three multivariate logistic regression models, these ORs changed little (Table 2). The fully adjusted ORs was 2.76 (95% CI, 1.53-4.96) for learning disability, 3.58 (95% CI, 1.92-6.55) for special education, and 0.77 (95% CI, 0.42-1.38) for ADHD, when comparing children with positive and negative anti-EBV. In gender-stratified analyses, the corresponding ORs for learning disability and special education were larger among males than among females [for learning disability, 2.99 (95% CI: 1.47-6.07) vs. 2.11 (95% CI: 0.79-5.62) and for special education, 5.42 (95% CI: 2.40-12.26) vs. 2.06 (95% CI: 0.83-5.10)]. However, there were no significant EBV-gender interactions in the analyses of learning disability and special education.

**Table 2.**
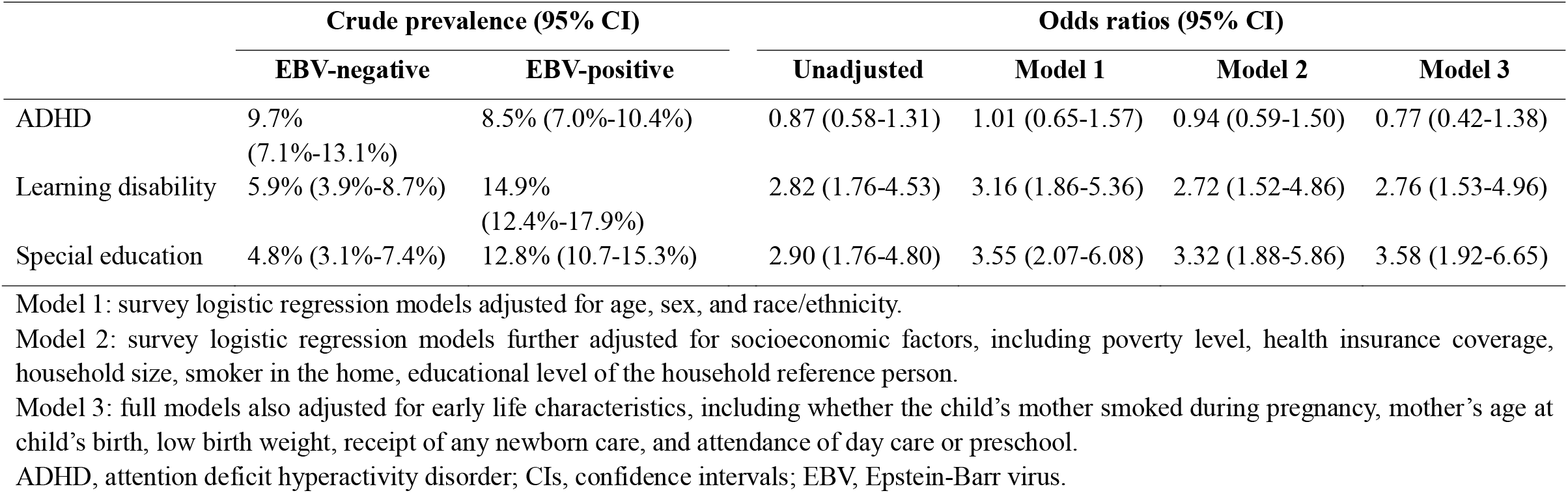
Relative Odds of Neurodevelopmental Outcomes among EBV-positive Children Compared with EBV-negative Children.

|  | Crude prevalence (95% CI) |  | Odds ratios (95% CI) |  |  |  |
| --- | --- | --- | --- | --- | --- | --- |
|  | EBV-negative | EBV-positive | Unadjusted | Model 1 | Model 2 | Model 3 |
| ADHD | 9.7%<br>(7.1%-13.1%) | 8.5% (7.0%-10.4%) | 0.87 (0.58-1.31) | 1.01 (0.65-1.57) | 0.94 (0.59-1.50) | 0.77 (0.42-1.38) |
| Learning disability | 5.9% (3.9%-8.7%) | 14.9%<br>(12.4%-17.9%) | 2.82 (1.76-4.53) | 3.16 (1.86-5.36) | 2.72 (1.52-4.86) | 2.76 (1.53-4.96) |
| Special education | 4.8% (3.1%-7.4%) | 12.8% (10.7-15.3%) | 2.90 (1.76-4.80) | 3.55 (2.07-6.08) | 3.32 (1.88-5.86) | 3.58 (1.92-6.65) |

Model 1: survey logistic regression models adjusted for age, sex, and race/ethnicity.
ADHD, attention deficit hyperactivity disorder; CIs, confidence intervals; EBV, Epstein-Barr virus.

## 4 Discussion

In the current study, based on a large representative sample, we first found that EBV seropositivity was associated with learning disability and special education among U.S. children. Using different models adjusting for potential confounding factors, these associations were strong and significant (Table 2). We found no association between EBV seropositivity and ADHD, which is consistent with previous studies (12, 15). As shown in Figure 1, learning disability and special education had much overlap in population. Thus, ORs for the two outcomes are likely to have the same direction. To our knowledge, no previous studies have evaluated the association of EBV with learning disability in children and only one small-sized case-control study evaluated the association with special education (16). The authors found no significant difference in EBV seroprevalence among children receiving special education (30/56) and their healthy peers (26/56) in Hampshire, England (16). Thus, the associations of EBV with learning disability and special education in our findings need replication.

EBV can cause injury to CNS (3-5). CNS complications occur in 1%-18% of patients with infectious mononucleosis and include meningitis, encephalitis, and psychiatric abnormalities (5). Especially in children, CNS complications can be the only clinical manifestations of EBV infection (17). Since the human CNS continues to develop throughout childhood and adolescence (18), infections of CNS during this period could increase the risk of neurodevelopment impairment. The impairment may be caused through direct CNS injury by pathogens, or through other pathways involving inflammation (8, 19). Other EBV-associated diseases are also linked to neurodevelopment impairment. For example, studies found that EBV seropositivity is more common in pediatric patients with multiple sclerosis than in their healthy peers and the former tends to have cognitive impairment (20-22).

Studies of general child populations failed to find a positive association of EBV with cognitive functioning (10-12). Khandaker *et al* found that early-life exposure to EBV was associated with lower, but not significant, IQ in adolescence from a population-based birth cohort in England (11). Karachaliou *et al* found no association of EBV seropositivity with cognitive development assessed with the McCarthy Scale of Children’s Abilities in 674 Greek children aged 4 years (12). Jonker *et al* also failed to find an association of EBV seropositivity at age 16 with immediate memory and executive functioning measured at age 18 in a Dutch adolescent population (10). But not like the current study, the three studies excluded children with neurodevelopmental conditions, while these children may have a learning disability and have the need for special education.

Our study has several strengths. The first is the representative sample of U.S. child population and a relatively large sample size. Second, the ORs for learning disability and special education are relatively high and consistent in different models adjusted for potential confounding factors. Third, EBV antibody was tested using a robust EIA. Equivocal results only occurred in approximately 0.5% of the cases and testing results were 96.7% concordant with those of the former IFA standard (13).

The main limitation of our study is the cross-sectional design, which is unable to draw causal inferences for the identified associations. EBV is spread by contact with oral secretions. Due to poor hand hygiene, children with a learning disability or receiving special education are expected to be exposed to EBV more likely (23). There are other mechanisms to explain the associations identified in our study. For example, EBV seropositivity is associated with susceptibility and severity of other infections in childhood, such as hand, foot, and mouth disease, of which severe cases may have neurodevelopmental sequelae (24). Thus, prospective studies to examine the associations identified in the current cross-sectional study are warranted.

The other limitation is that the assessment of ADHD, learning disability, and special education utilization was based on parental/guardian response. The assessment, especially for ADHD and learning disability, lacked a clear definition, and parental/guardian response may be not very reliable. In addition, ADHD and conditions with learning disabilities or needing special education present heterogeneous groups of neurodevelopmental abnormalities (25, 26). Future studies are suggested to collect data of high quality for neurodevelopmental outcomes.

Another limitation is that NHANES is not designed to answer a specific question related to health. The datasets are explored and analyzed repeatedly by a number of researchers from a wide range of disciplines. Publication bias exists as studies with statistically significant results are more likely to be published than those without (27). As a result, the actual statistical power of studies based on NHANES is lower than that of a planned study for a specific question. Nevertheless, this would be less likely to change the findings of associations of EBV with learning disability and special education in our analyses due to their statistically significant and high ORs.

## Data Availability

All data produced are available online at: https://wwwn.cdc.gov/nchs/nhanes/Default.aspx

## 5 Conclusions

In summary, we found that EBV seropositivity was associated with learning disability and special education among children. Therefore, if EBV causally associated with conditions of learning disability and needing special education in childhood, prevention and/or treatment strategies of EBV infection, such as hand hygiene, vaccination, or antiviral therapy, could serve to reduce related conditions in the general U.S. child population. Future studies that longitudinally examine the associations of EBV with learning disability and special education are warranted.

## 6 Data Availability Statement

Publicly available datasets were analyzed in this study. This data can be found here: https://wwwn.cdc.gov/nchs/nhanes/Default.aspx

## 8 Author Contributions

JW, YL, and XG conceptualized and designed the study, carried out the initial analyses, and drafted the initial manuscript. XZ and YX supervised data analyses and critically reviewed the manuscript. WW and YX conceptualized and designed the study, analyzed data, and reviewed and revised the manuscript. All authors have read and approved the final manuscript.

## Reference

1. de-Thé G, Day NE, Geser A, Lavoué MF, Ho JH, Simons MJ, et al. Sero-epidemiology of the Epstein-Barr virus: preliminary analysis of an international study - a review. IARC scientific publications (1975) (11 Pt 2):3–16. Epub 1975/01/01. PubMed PMID: 191375.

2. Cohen JI. Epstein-Barr virus infection. The New England journal of medicine (2000) 343(7):481–92. Epub 2000/08/17. doi: 10.1056/nejm200008173430707. PubMed PMID: 10944566.

3. Sumaya CV, Ench Y. Epstein-Barr virus infectious mononucleosis in children. I. Clinical and general laboratory findings. Pediatrics (1985) 75(6):1003–10. Epub 1985/06/01. PubMed PMID: 2987784.

4. Tsai MH, Hsu CY, Yen MH, Yan DC, Chiu CH, Huang YC, et al. Epstein-Barr virus-associated infectious mononucleosis and risk factor analysis for complications in hospitalized children. Journal of microbiology, immunology, and infection = Wei mian yu gan ran za zhi (2005) 38(4):255–61. Epub 2005/08/25. PubMed PMID: 16118672.

5. Weinberg A, Li S, Palmer M, Tyler KL. Quantitative CSF PCR in Epstein-Barr virus infections of the central nervous system. Annals of neurology (2002) 52(5):543–8. Epub 2002/10/29. doi: 10.1002/ana.10321. PubMed PMID: 12402250.

6. Coghill AE, Hildesheim A. Epstein-Barr virus antibodies and the risk of associated malignancies: review of the literature. American journal of epidemiology (2014) 180(7):687–95. Epub 2014/08/30. doi: 10.1093/aje/kwu176. PubMed PMID: 25167864; PubMed Central PMCID: PMCPMC4271109.

7. Houen G, Trier NH. Epstein-Barr Virus and Systemic Autoimmune Diseases. Frontiers in immunology (2020) 11:587380. Epub 2021/01/26. doi: 10.3389/fimmu.2020.587380. PubMed PMID: 33488588; PubMed Central PMCID: PMCPMC7817975.

8. John CC, Black MM, Nelson CA, 3rd. Neurodevelopment: The Impact of Nutrition and Inflammation During Early to Middle Childhood in Low-Resource Settings. Pediatrics (2017) 139(Suppl 1):S59–s71. Epub 2017/06/01. doi: 10.1542/peds.2016-2828H. PubMed PMID: 28562249; PubMed Central PMCID: PMCPMC5694688.

9. Kleinschmidt-DeMasters BK, Gilden DH. The expanding spectrum of herpesvirus infections of the nervous system. Brain pathology (Zurich, Switzerland) (2001) 11(4):440–51. Epub 2001/09/15. doi: 10.1111/j.1750-3639.2001.tb00413.x. PubMed PMID: 11556690.

10. Jonker I, Klein HC, Duivis HE, Yolken RH, Rosmalen JG, Schoevers RA. Association between exposure to HSV1 and cognitive functioning in a general population of adolescents. The TRAILS study. PloS one (2014) 9(7):e101549. Epub 2014/07/02. doi: 10.1371/journal.pone.0101549. PubMed PMID: 24983885; PubMed Central PMCID: PMCPMC4077793.

11. Khandaker GM, Stochl J, Zammit S, Lewis G, Jones PB. Childhood Epstein-Barr Virus infection and subsequent risk of psychotic experiences in adolescence: a population-based prospective serological study. Schizophrenia research (2014) 158(1-3):19–24. Epub 2014/07/23. doi: 10.1016/j.schres.2014.05.019. PubMed PMID: 25048425; PubMed Central PMCID: PMCPMC4561501.

12. Karachaliou M, Chatzi L, Roumeliotaki T, Kampouri M, Kyriklaki A, Koutra K, et al. Common infections with polyomaviruses and herpesviruses and neuropsychological development at 4 years of age, the Rhea birth cohort in Crete, Greece. Journal of child psychology and psychiatry, and allied disciplines (2016) 57(11):1268–76. Epub 2016/11/03. doi: 10.1111/jcpp.12582. PubMed PMID: 27334233.

13. Balfour HH, Jr., Sifakis F, Sliman JA, Knight JA, Schmeling DO, Thomas W. Age-specific prevalence of Epstein-Barr virus infection among individuals aged 6-19 years in the United States and factors affecting its acquisition. The Journal of infectious diseases (2013) 208(8):1286–93. Epub 2013/07/23. doi: 10.1093/infdis/jit321. PubMed PMID: 23868878.

14. Quiros-Alcala L, Mehta S, Eskenazi B. Pyrethroid pesticide exposure and parental report of learning disability and attention deficit/hyperactivity disorder in U.S. children: NHANES 1999-2002. Environ Health Perspect (2014) 122(12):1336–42. Epub 2014/09/06. doi: 10.1289/ehp.1308031. PubMed PMID: 25192380; PubMed Central PMCID: PMC4256700.

15. Bekdas M, Tufan AE, Hakyemez IN, Tas T, Altunhan H, Demircioglu F, et al. Subclinical immune reactions to viral infections may correlate with child and adolescent diagnosis of attention-deficit/hyperactivity disorder: a preliminary study from Turkey. African health sciences (2014) 14(2):439–45. Epub 2014/10/17. doi: 10.4314/ahs.v14i2.21. PubMed PMID: 25320595; PubMed Central PMCID: PMCPMC4196392.

16. Holme CO. Previous Epstein-Barr virus infections in children with special educational needs. J Infect (1982) 4(1):27–32. Epub 1982/01/01. doi: 10.1016/s0163-4453(82)90901-x. PubMed PMID: 6309975.

17. Kasper DL, Fauci AS. Harrison’s Infectious Diseases, 3/E: McGraw-Hill Education (2016).

18. de Graaf-Peters VB, Hadders-Algra M. Ontogeny of the human central nervous system: what is happening when? Early human development (2006) 82(4):257–66. Epub 2005/12/20. doi: 10.1016/j.earlhumdev.2005.10.013. PubMed PMID: 16360292.

19. Theoharides TC, Kavalioti M, Martinotti R. Factors adversely influencing neurodevelopment. Journal of biological regulators and homeostatic agents (2019) 33(6):1663–7. Epub 2020/01/14. doi: 10.23812/19-33n6Edit_Theoharides. PubMed PMID: 31928596.

20. Alotaibi S, Kennedy J, Tellier R, Stephens D, Banwell B. Epstein-Barr virus in pediatric multiple sclerosis. Jama (2004) 291(15):1875–9. Epub 2004/04/22. doi: 10.1001/jama.291.15.1875. PubMed PMID: 15100207.

21. Houen G, Trier NH, Frederiksen JL. Epstein-Barr Virus and Multiple Sclerosis. Frontiers in immunology (2020) 11:587078. Epub 2021/01/05. doi: 10.3389/fimmu.2020.587078. PubMed PMID: 33391262; PubMed Central PMCID: PMCPMC7773893.

22. Ekmekci O. Pediatric Multiple Sclerosis and Cognition: A Review of Clinical, Neuropsychologic, and Neuroradiologic Features. Behavioural neurology (2017) 2017:1463570. Epub 2018/02/13. doi: 10.1155/2017/1463570. PubMed PMID: 29434433; PubMed Central PMCID: PMCPMC5757108.

23. Lee RL, Leung C, Tong WK, Chen H, Lee PH. Comparative efficacy of a simplified handwashing program for improvement in hand hygiene and reduction of school absenteeism among children with intellectual disability. American journal of infection control (2015) 43(9):907–12. Epub 2016/07/09. doi: 10.1016/j.ajic.2015.03.023. PubMed PMID: 27387071.

24. Li Y, Dang S, Deng H, Wang W, Jia X, Gao N, et al. Breastfeeding, previous Epstein-Barr virus infection, Enterovirus 71 infection, and rural residence are associated with the severity of hand, foot, and mouth disease. European journal of pediatrics (2013) 172(5):661–6. Epub 2013/01/25. doi: 10.1007/s00431-013-1939-1. PubMed PMID: 23344210.

25. Handler SM, Fierson WM, Section on O. Learning disabilities, dyslexia, and vision. Pediatrics (2011) 127(3):e818–56. Epub 2011/03/02. doi: 10.1542/peds.2010-3670. PubMed PMID: 21357342.

26. Luo Y, Weibman D, Halperin JM, Li X. A Review of Heterogeneity in Attention Deficit/Hyperactivity Disorder (ADHD). Frontiers in human neuroscience (2019) 13:42. Epub 2019/02/26. doi: 10.3389/fnhum.2019.00042. PubMed PMID: 30804772; PubMed Central PMCID: PMCPMC6378275.

27. Easterbrook PJ, Berlin JA, Gopalan R, Matthews DR. Publication bias in clinical research. Lancet (London, England) (1991) 337(8746):867–72. Epub 1991/04/13. doi: 10.1016/0140-6736(91)90201-y. PubMed PMID: 1672966.

